# A Sanger-based approach for scaling up screening of SARS-CoV-2 variants of interest and concern

**DOI:** 10.1101/2021.03.20.21253956

**Authors:** Matheus Filgueira Bezerra, Lais Ceschini Machado, Viviane do Carmo Vasconcelos de Carvalho, Cássia Docena, Sinval Pinto Brandão-Filho, Constância Flávia Junqueira Ayres, Marcelo Henrique Santos Paiva, Gabriel Luz Wallau

## Abstract

The global spread of new SARS-CoV-2 variants of concern underscore an urgent need of simple deployed molecular tools that can differentiate these lineages. Several tools and protocols have been shared since the beginning of the COVID-19 pandemic, but they need to be timely adapted to cope with SARS-CoV-2 evolution. Although whole-genome sequencing (WGS) of the virus genetic material have been widely used, it still presents practical difficulties such as high cost, shortage of available reagents in the global market, need of a specialized laboratorial infrastructure and well-trained staff. These limitations result in genomic surveillance blackouts across several countries. Here we propose a rapid and accessible protocol based on Sanger sequencing of a single PCR fragment that is able to identify and discriminate all SARS-CoV-2 variants of concern (VOCs) identified so far, according to each characteristic mutational profile at the Spike-RBD region (K417N/T, E484K, N501Y, A570D). Twelve COVID-19 samples from Brazilian patients were evaluated for both WGS and Sanger sequencing: three from P.2, two from P.1 and seven from B.1.1 lineage. All results from the Sanger sequencing method perfectly matched the mutational profile of VOCs and non-VOCs described by WGS. In summary, this approach allows a much broader network of laboratories to perform molecular surveillance of SARS-CoV-2 VOCs and report results within a shorter time frame, which is of utmost importance in the context of rapid public health decisions in a fast evolving worldwide pandemic.

---

As of December 2020, the United Kingdom reported a new SARS-CoV-2 variant, the B.1.1.7 lineage, which presented a higher transmissibility rate, bringing deep concerns about the prospects of the COVID-19 pandemic (1). Shortly after, other so-called “Variants Of Concern” (VOCs) were reported in South Africa (B.1.3.51), Brazil (P.1) and more recently, in the U.S.A (B.1.526) (2-4). Specific mutations, such as the N501Y and the E484K, in the residue binding domain (RBD) of the Spike protein are recurrent across the VOCs. These mutations play an important role on the lineage phenotype, allowing higher affinity to the human ACE2 receptor and/or immune evasion from previously elicited antibodies (5,6). It is likely that continuous circulation of SARS-CoV-2 in previously exposed and vaccinee populations will drive SARS-CoV-2 evolution towards lineages with increased transmissibility and escape from immune responses, allowing these variants to spread quickly throughout the world (6,7). In this scenario, the development of large-scale molecular surveillance strategies to monitor SARS-CoV-2 VOCs is crucial to provide timely information for proper public health control and adaptation of vaccination measures.

Since the release of the first SARS-CoV-2 genome, many molecular tools have been adapted to detect and monitor this virus in parallel with its emerging genomic changes (8). One of the most employed tools, capable of yielding unprecedented results is the whole genome sequencing (WGS) of SARS-CoV-2 from clinical samples. However, WGS is still very expensive to be applied as a front-line method for massive testing, particularly in underdeveloped and developing countries. Additionally, other PCR-based methodologies have been developed as well, focusing mainly on lineage-specific deletions of emerging VOCs and/or Spike mutation differentiation based on amplification dropouts and specific probes in RT-PCR assays (9,10). However, worldwide shortage of imported reagents, limited laboratorial infrastructure and the need of well-trained staff are other limitations commonly faced by these molecular protocols, resulting in surveillance blackouts in many countries. To illustrate the large discrepancies in genomic surveillance data observed during the Covid-19 pandemic, whilst 6.5% (270,762/4.1 × 10^6^) of the UK confirmed cases had their genomes sequenced, only 0.03% (3,430/10.5 × 10^6^) of the Brazilian confirmed cases were sequenced by early March (11). Therefore, the establishment and standardization of as many molecular protocols as possible that help to scale up the SARS-CoV-2 VOCs screening is highly desirable. Here we propose a rapid and accessible protocol based on Sanger sequencing that is able to identify and discriminate SARS-CoV-2 VOCs, according to each characteristic mutational profile at the Spike-RBD region.

In order to access whether the amplicon used in this study is able to cover key SARS-CoV-2 mutations, we accessed Twelve COVID-19 positive samples (RT-PCR - Ct values below 25) derived from symptomatic patients of both Pernambuco (Northeast Brazil) and Amazonas (North Brazil) states that had been previously genomic sequenced (8). The study was approved by the local Ethical Committee (CAAE32333120.4.0000.5190). RNA extractions were performed in a BSL-3 facility laboratory with a robotic platform using the Maxwell® 16 Viral Total Nucleic Acid Purification Kit (Promega, Wisconsin-USA), following the manufacturer’s instructions. The molecular diagnosis of SARS-CoV-2 was performed using the Kit Molecular BioManguinhos SARS-CoV-2 (E/RP).

High Capacity cDNA Reverse-Transcription kit (Applied Biosystems) was used for reverse transcription, following the manufacturer’s instruction. Next, cDNA was subjected to PCR with Platinum Taq-polymerase (Invitrogen) and primers flanking the regions between the nucleotide positions 22797 and 23522 of the Wuhan (Wu-1) reference genome, covering key amino acid replacements commonly found in VOCs RBD domain of the Spike protein (76 Left: 5’-AGGGCAAACTGGAAAGATTGCT-3’ and 77 Right: 5’-CAGCCCCTATTAAACAGCCTGC-3’ designed by https://www.protocols.io/view/ncov-2019-sequencing-protocol-bbmuik6w). PCR conditions were: 98 °C for 5 minutes s, 98°C for 30 seconds, 59°C for 30 seconds and 72°C for 45 seconds during 35 cycles and final extension of 5 min at 72°C. Primer and magnesium chloride concentrations in the PCR were 0.2 µM and 1 mM, respectively. Amplified PCR products were verified in a 1.5% Agarose gel stained with Sybr Safe (Sigma-Aldrich), quantified in a NanoDrop OneC Microvolume UV-Vis Spectrophotometer (Thermo-Fischer, USA) and diluted to 30 ng/uL. Sequencing reactions were performed with BigDye Terminator v3.1 (Applied Biosystems) and ran in capillary electrophoresis (ABI 3500, Applied Biosystems). Contigs from forward and reverse strands were built and analyzed using the CodonCode aligner v3.7.1 software and figures were built using the Biorender platform. Samples were assigned to a lineage according to the mutational profile (Table 1).

**Table 1.** Sars cov-2 lineages according to the mutational profile in Sanger sequencing.

| Lineage | First report | Mutation |  |  |  |  |  |  |
| --- | --- | --- | --- | --- | --- | --- | --- | --- |
|  |  | K417N | K417T | L452R | S477N | E484K | N501Y | A570 |
| **P.1** | Brazil (Amazon) | - | present | - | - | present | present | - |
| **P.2** | Brazil | - | - | - | - | present | - | - |
| **B1.1.7** | U.K | - | - | - | - | - | present | present |
| **B.1.3.51** | South Africa | present | - | - | - | present | present | - |
| **CAL.20C** | U.S.A (California) | - | - | present | - | - | - | - |
| **B.1.526** | U.S.A (New York) | - | - | - | present | present | - | - |

According to the WGS, from the twelve COVID-19 samples evaluated, seven were from the B.1.1 lineage (non-VOC), three were P.2 and two were P.1. Remarkably, in a blind comparison to WGS (gold standard), all results from the Sanger sequencing method matched those from WGS method. The K417, E484 and N501Y mutations were identified in the P.1 cases and the E484K (in absence of the others) in the P.2 cases (Table 1).

Within the sequencing of a single 725 base pairs PCR fragment (Figure 1), this approach could successfully detect VOC-associated mutations and correctly classify samples according to the WGS data. Moreover, the flanked region also covers other relevant circulating RBD mutations (Figure 1) and potentially, new mutations that have not been identified yet. Together, these features overcome some of the limitations of allelic-specific PCR methods, such as the need of one specific probe or primer for each mutation to be evaluated and previous knowledge of the circulating mutations (10). Furthermore, high-quality electropherograms were obtained without a PCR purification step, reducing costs and time of sample processing, which is particularly useful for large-scale application of the method. Another advantage of this approach is that primers can be easily adjusted without major protocol modifications, in case newly described mutations need to be detected. On the other hand, it is important to highlight that Sanger sequencing is normally more time consuming than allelic-specific RT-PCR and hence with a comparative reduced scaling capacity, but it brings some advantages such as more genetic data that helps to tease apart different VOCs and the possibility of detecting new emerging RBD mutations.

**Figure 1.**
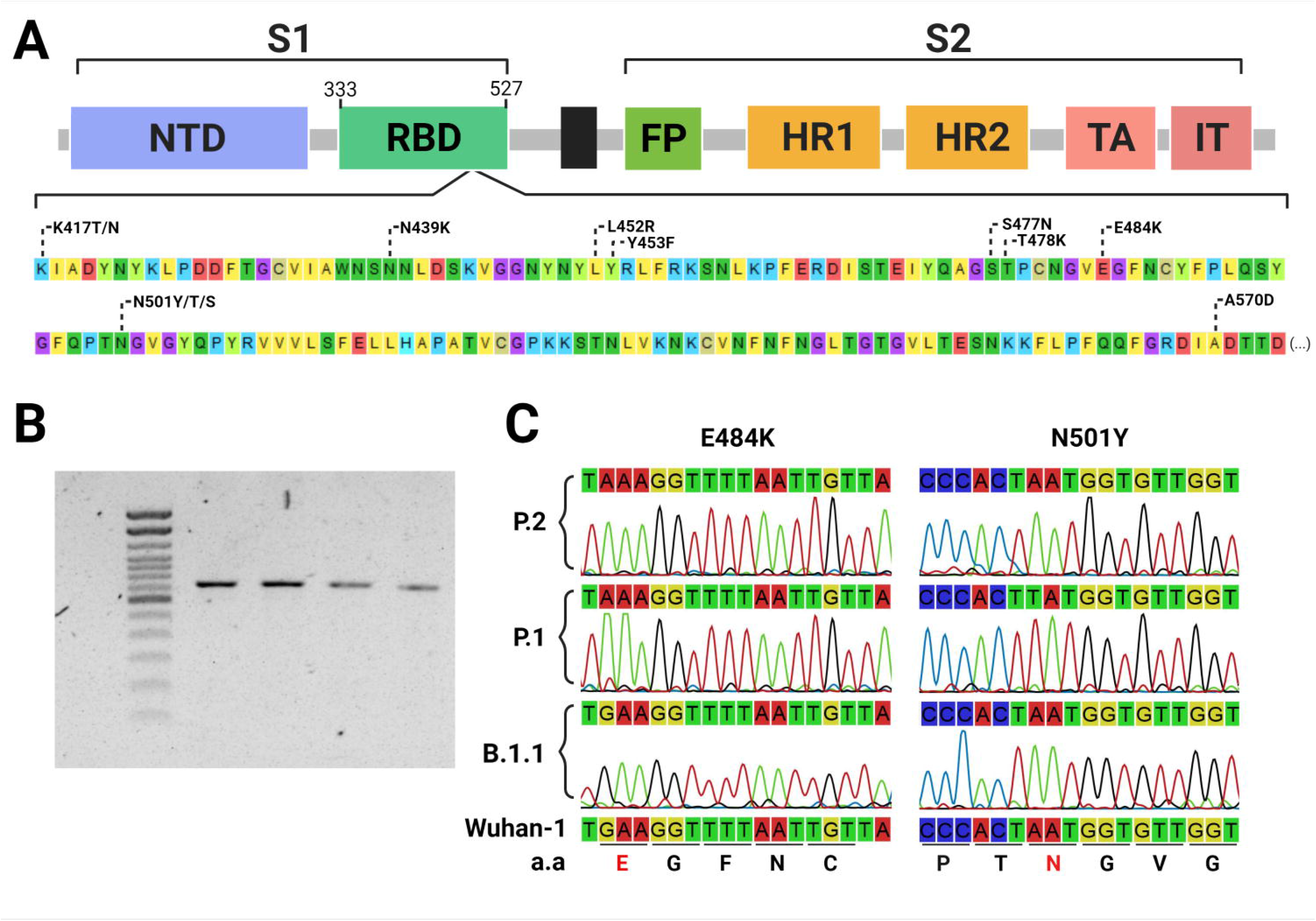
Identification of Sars CoV-2 Spike-RBD mutations using Sanger sequencing. Commonly found RBD mutations flanked by the primer set (nucleotide positions from 22797 to 23522 at the Wu-1 genome) used for sequencing, including key mutations to enable identifying variants of concern and interest **(A)**. 725 bp PCR fragments amplified from Sars Cov-2 cDNA **(B)**. Sections from the eletropherograms obtained by Sanger sequencing showing the E484K and N501Y VOC-associated mutations **(C)**.

It is important to highlight that this approach does not substitute WGS and other PCR-based assays and could be used in combination to further validate the VOCs results mainly with WGS to uncover other important mutation at the SAR-CoV-2 genome, but it will allows a much broader network of laboratories to perform molecular surveillance of SARS-CoV-2 VOCs, reporting results within a shorter time frame and in larger amounts, which is of utmost importance in the context of rapid public health decisions in a fast evolving worldwide pandemic.

## Data Availability

All genomes generated in this study are deposited on GISAID under the accessions: EPI_ISL_500460, EPI_ISL_500461, EPI_ISL_500865, EPI_ISL_500868, EPI_ISL_500872, EPI_ISL_500477, EPI_ISL_500482, EPI_ISL_1239012, EPI_ISL_1239013, EPI_ISL_1239014, EPI_ISL_1239015, EPI_ISL_1239016

https://www.gisaid.org

## ACKNOWLEDGMENTS

We would like to thank the COVID-IAM and LACEN-PE teams for providing the samples to sequence the SARS-CoV-2 genomes, the Technological Platform Core and the Bioinformatic Core of the Aggeu Magalhaes Institute for the support with their research facilities.

## AUTHOR CONTRIBUTIONS

M.F.B conceived the study, performed experiments, collected/analyzed data and drafted the manuscript. L.C.M, V.C.V.C and C.D performed experiments. S.P.B.F and C.F.J.A obtained patient samples, updated the clinical data and corrected the manuscript. M.H.S.P and G.L.W conceived and designed the study, analyzed data and gave the final approval of the version to be submitted.

